# The implications of using maternity care deserts to measure progress in access to obstetric care: A mixed-integer optimization analysis

**DOI:** 10.1101/2023.10.31.23297779

**Authors:** Meghan E. Meredith, Lauren N. Steimle, Stephanie M. Radke

## Abstract

**Background:** Lack of access to risk-appropriate maternity services, particularly for rural residents, is thought to be a leading contributor to disparities in maternal morbidity and mortality. There are several existing measures of access to obstetric care in the literature and popular media. In this study, we explored how current measures of obstetric access inform the number and location of additional obstetric care facilities required to improve access.

**Methods:** We formulated two facility location optimization models to determine the number of new facilities required to minimize the number of reproductive-aged women who lack access to obstetric care. We define regions with a lack of access as either maternity care deserts, designated by the March of Dimes to be counties with no obstetric care facility or obstetric providers, or regions further than 50 miles from critical care obstetric (CCO) services. We gathered information on hospitals with obstetric services from Georgia Department of Public Health public reports and estimated the female reproductive-age population by census block group using the American Community Survey.

**Results:** Out of the 1,910,308 reproductive-aged women who live in Georgia, 104,158 (5.5%) live in maternity care deserts, 150,563 (7.9%) reproductive-aged women live further than 50 miles from CCO services, and 38,202 (2.0%) live in both maternity care desert and further than 50 miles from CCO services. Our optimization analysis suggests that at least 56 new obstetric care facilities (a 67% increase) would be required to eliminate maternity care deserts in Georgia. However, and the expansion of 8 facilities would ensure all women in Georgia live within 50 miles of CCO services.

**Conclusions:** Current measures of access to obstetric care may not be sufficient for evaluating access and planning action toward improvements. In a state like Georgia with a large number of small counties, eliminating maternity care deserts would require a prohibitively large number of new obstetric care facilities. This work suggests that additional measures and tools are needed to estimate the number and type of obstetric care facilities that best match practical resources to meet obstetric care needs.

## 1. Background

The maternal mortality rate in the United States (U.S.), 32.9 deaths per 100,000 live births as of 2021, is the highest among developed countries and has increased by 89% since 2018.^1,2^ There is evidence that upwards of 80% of maternal deaths in the U.S. are preventable.^3^ Among the factors contributing to the maternal mortality crisis in the U.S. is a lack of access to risk-appropriate care and an undersupply of maternal healthcare providers.^2^

Rural access to obstetric services has been declining in recent years. Over half of rural counties did not have a facility offering obstetric services in 2014, and this number grew by 2.7% from 2014 to 2018.^4^ Administrators cite financial concerns, shortages of obstetric professionals, and low volume as reasons for closing their obstetric units.^5,6^ Lack of access to obstetric services is associated with adverse maternal outcomes, adverse neonatal outcomes, and prenatal stress.^7–11^ Recent findings suggest a lack of access and disparities in geographic access will persist unless facility-level infrastructure is expanded.^12^ However, geographic access to obstetric care is measured in several ways, which causes uncertainty about how to optimally invest in infrastructure to expand access. One common measure of access in the academic literature and news media is the maternity care desert, as defined by the March of Dimes.^13,14^ The March of Dimes categorizes counties with a lack of access to care (no hospital or birth center offering obstetric care and no obstetric providers) as maternity care deserts. As of 2022, more than 2.2 million reproductive-aged women in the U.S. live in maternity care deserts.^15^ Studies have shown that pregnant women who live in maternity care deserts have higher rates of infant and maternal mortality.^16,17^ However, the maternity care deserts access measure does not necessarily reflect distance to care because counties differ in size and some pregnant women within a county may live close to an obstetric facility in a neighboring county. Other studies have measured geographic access as driving time to the nearest facility offering obstetric services at different levels of care^12,18^ and distance to the nearest facility offering critical care obstetric (CCO) services^19,20^ as key measures for quantifying potential access.

In contrast to these existing studies that measure current levels of access, we considered the implications of using these metrics as key performance indicators for tracking improvements in access to obstetric care. In particular, we asked: what is required for states to reduce the number of women who lack access to obstetric care, as defined by two different access to care measures? To answer this question, we considered the implications of expanding access to care through facility expansions by drawing upon *mathematical optimization*. Optimization is a mathematical science that is widely used to identify the ideal solution while considering the complex interactions and constraints within a system.^21^ The specific type of optimization modeling framework, facility location modeling, has often been used to evaluate the ideal placement of healthcare facilities to ensure proper coverage of a patient population.^22–24^ A comprehensive review of healthcare facility location modeling is provided by Admadi-Javid et al.^25^

In this article, we characterized access to obstetric care using existing access measures and evaluated these existing measures by determining how many facilities are needed to provide a sufficient level of access according to these measures. We focused on the State of Georgia because Georgia has one of the highest rates of maternal mortality in the U.S. – almost twice as high as the national rate.^26^ As of 2019 more than 75% of Georgia’s 159 counties had no hospital or birth center offering obstetric care.^15^ Georgia does have a set of Regional Perinatal Centers whose mission is to coordinate access to optimal and risk-appropriate maternal and infant care.^27^ Georgia is taking multiple initiatives to improve obstetric outcomes, including extending Medicaid coverage, introducing quality improvement initiatives, verifying levels of maternal care in Georgia hospitals, and expanding home visiting in rural counties.^28^

First, we characterized regions that lack access to obstetric care using two commonly used measures in the literature: (1) the March of Dimes maternity care desert measure^15^ and (2) regions that are further than 50 miles from the closest facility that provides CCO services. Upon defining a region as lacking access or not, we reported the total number of reproductive-aged women who lack access to obstetric care according to each measure. Finally, we analyzed how many facilities would be needed in the state of Georgia to reduce the number of reproductive-aged women who lack access to obstetric care by 50% and 100%.

The goal of this study is to characterize regions defined to have a lack of access to obstetric care based on two existing measures of access and to determine the facility interventions required to improve access according to these measures. We hypothesized that obstetric facility expansion policies focused on reducing maternity care deserts alone are impractical and could have negative consequences and policies focusing on reducing distance to CCO services alone are not aligned with risk-appropriate care for the majority of pregnancies, revealing the need for new measures of geographic access to high-quality, risk-appropriate care which can be used as targets for policy intervention.

## 2. Methods

### 2.1 Data Sources

First, we collected data to infer the geographic distribution of obstetric healthcare facilities and providers, as well as the geographic distribution of subpopulations and communities that would demand obstetric services. The data sources used are described below.

#### 2.1.1 Location of Facilities Providing Obstetric Care

We included obstetric facilities in Georgia that are classified as birth centers, or Perinatal Care Level 1, 2, or 3 hospitals according to the public records from Georgia’s Department of Public Health from 2017.^27^ The address of each obstetric facility was verified by the study team by cross-referencing with Google Maps, and the latitude and longitude of each obstetric facility were located using Python’s geopy package.^29^

#### 2.1.3 Location of Demand for Obstetric Care

To estimate the demand for obstetric care access, we used data from the American Community Survey (ACS) which provides population estimates for age and sex groups. We used the 2017 ACS 5-year estimates of the population of reproductive-aged women (18-44) in each census block group, which we assumed is proportional to the demand for obstetric care in each block group. We used 5-year estimates because they are the most reliable and they are collected for all small geographies including census block groups. To estimate the location of this demand, we used the latitude and longitude of center of population of each census block group as reported by the U.S. Census Bureau in 2010 to be consistent with our facility and population estimates data from 2017.^30^

#### 2.1.2 Distance to Obstetric Care

We calculated the distance between each obstetric facility and each obstetric care demand point using Great Circle distance^29^ in miles between the coordinates of each facility and each census block group center of population. Great Circle distance is the direct distance between two points accounting for the curvature of the earth, and is commonly used to estimate access to healthcare.^31,32^

### 2.2 Measures of Obstetric Access

We then determined which census block groups lack access to obstetric care according to the measures outlined below.

#### 2.2.1 Maternity Care Desert

We considered the March of Dimes definition of a maternity care desert which is defined to be a county that has zero hospitals or birth centers offering obstetric services and zero obstetric providers.^15^ Because maternity care deserts are defined at the county level and the distance measure is defined at the census block group level, we deemed any census block group in a maternity care desert county to be a maternity care desert census block group. Our study team validated Georgia maternity care deserts based on our data against the March of Dimes maternity care deserts dashboard and found they were consistent.^33^ We then used the following evaluation measures to compare these definitions.

#### 2.2.2 Distance to Critical Care Obstetric (CCO) Hospital

We evaluated the distance from the center of population of each census block group to its nearest facility offering CCO services. In line with previous studies,^20^ we characterized hospitals as offering CCO services if they are designated as Perinatal Care Level 3 obstetric hospitals. We refer to birth centers and Level 1 and 2 obstetric hospitals collectively as “lower-level” hospitals. These lower-level hospitals provide basic obstetric care but do not provide CCO services. We referred to public reporting from Georgia’s Department of Public Health to characterize each hospital’s level of care.^27^ We then evaluated whether the census block group population center is within the pre-specified distance threshold of 50 miles. A 50-mile threshold is commonly used because it approximates the farthest distance most people appear willing to travel for specialized medical care and it estimates the widely accepted “Golden Hour”. The “Golden Hour” stems from trauma care, where it is thought that critically injured patients have better outcomes if they receive definitive care within an hour of their injuries.^34^ This 50-mile threshold has been commonly used to estimate access to obstetric care,^19,20^ although it has not been validated for obstetric care.^35,36^

### 2.3 Evaluation Metrics

Using the measures above, we characterized each census block group as either having access to obstetric care or lacking access to obstetric care.

#### 2.3.1 Characterization of lack of access to obstetric care

First, we characterized the number of census block groups that lacked access to obstetric care according to different measures of access (i.e., maternity care desert, > 50 miles from CCO services, and both a maternity care desert and > 50 miles from CCO services). Additionally, we characterized the demographics of the populations within the census block groups that lacked access to obstetric care according to different measures of access.

#### 2.3.2 Other measures of access to obstetric care

We characterized the distribution of distance to the closest obstetric facility for different measures of access to obstetric care. We further characterized distance to care by level of care, calculating the distance to the closest facility offering Level 1, 2, and 3 care.

#### 2.3.3 Evaluating the need for facility expansion to improve access

We considered how many new facilities would hypothetically be needed to reduce the number of reproductive-aged women who lack access to obstetric care by 50% and 100%. To do so, we use a *mathematical optimization* model drawing from the facility location literature (see Appendix). This optimization model determined the optimal placement of new obstetric facilities to minimize the number of reproductive-aged women living in deserts. This model unrealistically assumed that we could readily build obstetric facilities anywhere we wanted. We revisit this assumption in the discussion.

We considered both measures of access to obstetric care in our optimization models. First, we investigated the number of new obstetric facilities that would hypothetically be required to reduce the number of women in maternity care deserts by a given percentage. To do so, we formulated a mathematical optimization model that minimized the total number of reproductive-aged women who live in maternity care deserts by introducing at most X new obstetric hospitals. This model returned the optimal location of these X new facilities. Here, X is a parameter that was varied to analyze the change in the number of reproductive-aged women living in maternity care deserts as more facilities are introduced. We also investigated the number of existing lower-level obstetric facilities that would need to be upgraded to provide CCO services to reduce the number of women living further than 50 miles from a CCO facility by a given percentage. We formulated a second mathematical optimization model that minimized the total number of reproductive-aged women living further than 50 miles from CCO services by optimally choosing at most X existing lower-level obstetric hospitals to upgrade to CCO.

## 3. Results

### 3.1. Characterization of lack of access to obstetric care

Figure 1 shows the regions that lack access to obstetric care according to the two access measures. In Georgia, 83 hospitals offer obstetric services. 56 counties are deemed to be maternity care deserts, which contain a combined 524 census blocks. In comparison, 650 census block groups from 53 counties are further than 50 miles to CCO services Table 1 shows that out of the 1,910,308 reproductive-aged women who live in Georgia, 104,158 (5.5%) live in maternity care deserts, 150,563 reproductive-aged women (7.9%) live more than (>) 50 miles from CCO services, and 38,202 (2.0%) live in both maternity care deserts and > 50 miles from CCO services.

**Figure 1.**
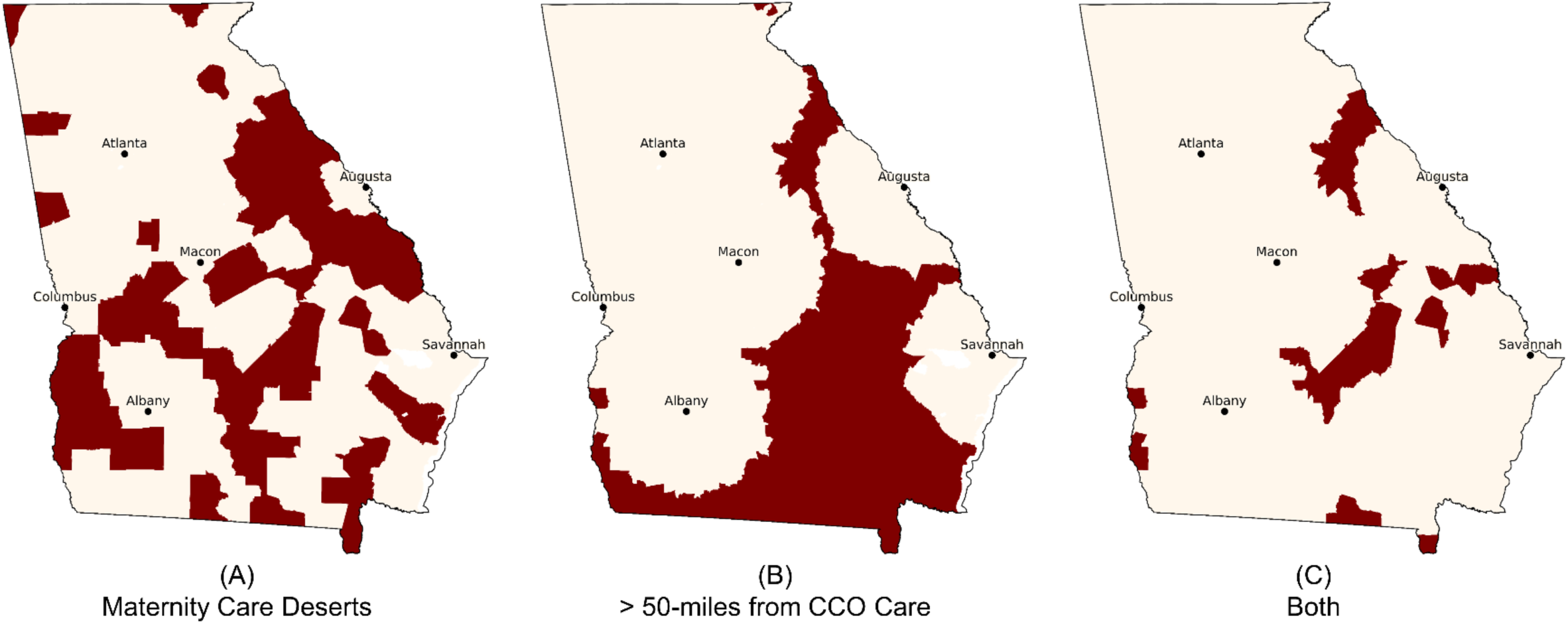
Current state of lack of access to obstetric care in Georgia under different definitions. (A) Maternity Care Deserts, (B) > 50 miles from CCO services, (C) both Maternity Care Deserts and > 50 miles from CCO services

**Table 1.** The characteristics of all people who live in Georgia by obstetric access and the ages of reproductive-aged females by obstetric access.

| **Characteristics of Georgia Population by Obstetric Access** |  |  |  |  |
| --- | --- | --- | --- | --- |
|  | Georgia Overall | Maternity Care Desert | > 50 miles from CCO Services | Maternity Care Desert & > 50 mi from CCO Services |
| **Total Population** | **10,201,635 (100.0%)** | **670,558 (6.6%)** | **890,237 (8.7%)** | **247,074 (2.4%)** |
| **Race** |  |  |  |  |
| White | 6,061,821 (59.4%) | 427,994 (63.8%) | 585,792 (65.8%) | 164,592 (66.6%) |
| Black/African American | 3,195,268 (31.3%) | 210,003 (31.3%) | 255,866 (28.7%) | 71,646 (29.0%) |
| American Indian/Alaska Native | 30,552 (0.3%) | 1,540 (0.2%) | 2,583 (0.3%) | 712 (0.3%) |
| Asian | 388,946 (3.8%) | 4,031 (0.6%) | 7,872 (0.9%) | 1,180 (0.5%) |
| Native Hawaiian/Pacific Islander | 5,237 (0.1%) | 569 (0.1%) | 264 (0.0%) | 54 (0.0%) |
| Other | 282,570 (2.8%) | 16,151 (2.4%) | 21,521 (2.4%) | 5,600 (2.3%) |
| Multiracial | 237,241 (2.3%) | 10,270 (1.5%) | 16,339 (1.8%) | 3,290 (1.3%) |
| **Ethnicity** |  |  |  |  |
| Hispanic/Latino | 950,380 (9.3%) | 37,438 (5.6%) | 57,444 (6.5%) | 16,797 (6.8%) |
| **Insurance** |  |  |  |  |
| No Insurance | 1,481,625 (14.8%) | 108,443 (16.9%) | 146,234 (17.2%) | 43,264 (18.4%) |
| Medicaid | 1,491,181 (14.9%) | 135,480 (21.1%) | 173,232 (20.4%) | 53,606 (22.8%) |
| **Poverty** | 1,679,030 (16.9%) | 150,938 (23.7%) | 198,171 (23.4%) | 58,789 (25.1%) |
| **Total Female Reproductive-aged (18-44)** | **1,910,308 (18.7%)** | **104,158 (15.5%)** | **150,563 (16.9%)** | **38,202 (15.5%)** |
| 18-24 | 492,292 (25.8%) | 27,149 (26.1%) | 41,512 (27.6%) | 9,662 (25.3%) |
| 25-34 | 709,387 (37.1%) | 37,555 (36.1%) | 55,886 (37.1%) | 13,856 (36.3%) |
| 35-44 | 708,629 (37.1%) | 39,454 (37.9%) | 53,165 (35.3%) | 14,684 (38.4%) |

In Georgia, 14.8% of people do not have insurance and 14.9% of people have Medicaid. These proportions are higher for people who live in regions characterized as maternity care deserts (16.9%, 21.1%), > 50 miles from CCO services (17.2%, 20.4%), and regions designated as both (18.4%, 22.8%). Also, in Georgia, 16.9% of people have an income below the federal poverty line. This proportion is higher in regions characterized as maternity care deserts (23.7%), > 50 miles from CCO services (23.4%), and regions designated as both (25.1%).

### 3.2 Other measures of access to obstetric care

Table 2 shows the number of reproductive-aged women who live within the specified distance from obstetric services for each level of care. Of the 104,158 reproductive-aged women who live in maternity care deserts, 63% are within 50 miles of CCO services, 97% are within 50 miles of Level 2 care, and 100% are within 50 miles of any obstetric care facility. Of the 150,563 reproductive-aged women who live > 50 miles from CCO services, 98% are within 50 miles of Level 2 care, 100% are within 50 miles of any obstetric care facility, and 75% do not live in a maternity care desert. Of the 1,806,150 reproductive-aged women who do not live in maternity care deserts, 93% are within 50 miles of CCO services. Similarly, of the 1,759,745 women who are within 50 miles of CCO services, 96% live in a county with an obstetric care facility.

**Table 2.** The number and proportion of the reproductive-aged women by obstetric access who live within the specified distance threshold of each level of obstetric care.

| **Reproductive-Aged Women Who Live Within Distance of Obstetric Care, N (%)** |  |  |  |  |  |  |
| --- | --- | --- | --- | --- | --- | --- |
| **Distance from Obstetric Care** |  | Georgia Overall | Maternity Care Deserts | Not in Maternity Care Deserts | > 50 miles from CCO services | ≤ 50 miles from CCO services |
| **Distance** | **Level of Care** | **N = 1,910,308**<br>**(100%)** | **N = 104,158**<br>**(5.5%)** | **N = 1,806,150**<br>**(94.5%)** | **N = 150,563**<br>**(7.9%)** | **N = 1,759,745**<br>**(92.1%)** |
| 25 miles | 1 | 1,194,235 (62%) | 41,860 (40%) | 1,152,375 (63%) | 80,529 (53%) | 1,113,706 (63%) |
|  | 2 | 1,546,787 (80%) | 54,577 (52%) | 1,492,210 (82%) | 86,879 (57%) | 1,459,908 (82%) |
|  | 3 | 1,490,107 (78%) | 17,789 (17%) | 1,472,318 (81%) | 0 (0%) | 1,490,107 (84%) |
|  | Any | 1,883,936 (98%) | 85,763 (82%) | 1,798,173 (99%) | 140,714 (93%) | 1,743,222 (99%) |
| 50 miles | 1 | 1,791,838 (93%) | 86,719 (83%) | 1,705,119 (94%) | 132,182 (87%) | 1,659,656 (94%) |
|  | 2 | 1,898,528 (99%) | 101,364 (97%) | 1,797,164 (99%) | 148,257 (98%) | 1,750,271 (99%) |
|  | 3 | 1,759,745 (92%) | 65,956 (63%) | 1,693,789 (93%) | 0 (0%) | 1,759,745 (100%) |
|  | Any | 1,910,308 (100%) | 104,158 (100%) | 1,806,150 (100%) | 150,563 (100%) | 1,759,745 (100%) |
| 100 miles | 1 | 1,910,308 (100%) | 104,158 (100%) | 1,806,150 (100%) | 150,563 (100%) | 1,759,745 (100%) |
|  | 2 | 1,910,308 (100%) | 104,158 (100%) | 1,806,150 (100%) | 150,563 (100%) | 1,759,745 (100%) |
|  | 3 | 1,909,715 (99%) | 103,630 (99%) | 1,806,085 (99%) | 149,970 (99%) | 1,759,745 (100%) |
|  | Any | 1,910,308 (100%) | 104,158 (100%) | 1,806,150 (100%) | 150,563 (100%) | 1,759,745 (100%) |
| **Not in Maternity Care Deserts** |  | 1,806,150 (95%) | 0 (0%) | 1,806,150 (100%) | 112,361 (75%) | 1,693,789 (96%) |

### 3.3 Responsiveness to interventions

Figure 2 shows the results of our optimization analysis. To hypothetically reduce the number of reproductive-aged women living in maternity care deserts by at least 50%, 16 new obstetric hospitals would be required in counties that are currently maternity care deserts. This would be an increase of 19% over the 83 current number of facilities offering obstetric services and would reduce the number of reproductive-aged women living in maternity care deserts from 104,158 to 51,477. To eliminate maternity care deserts in Georgia, 56 new obstetric hospitals would be required (a 67% increase in obstetric facilities; one facility for each county that is currently a maternity care desert).

**Figure 2.**
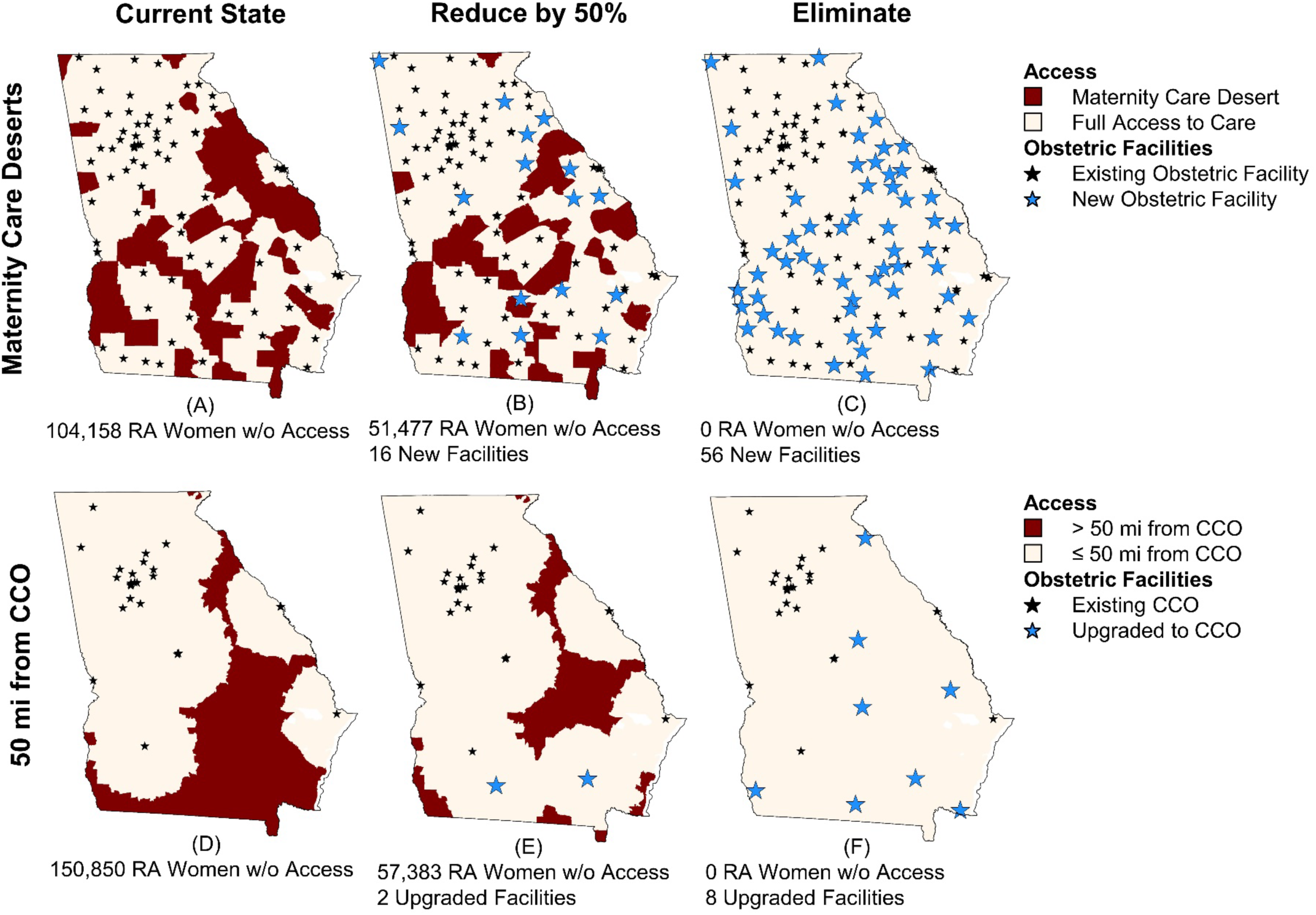
The number of obstetric facilities needed to be expanded to reduce the number of reproductive-aged (RA) women who lack access to obstetric care by 50% and 100% according to two measures of access.

Our optimization analysis shows that to reduce the number of reproductive-aged women living 50 miles from CCO services by at least 50% (from 150,563 to 57,338 reproductive-aged women) it would require upgrading 2 obstetric facilities to offer CCO services. To eliminate all census block groups that are > 50 miles from CCO services, a minimum of 8 facilities would need to be upgraded to offer CCO services.

Figure 3 shows how many facilities are needed to reduce the number of reproductive-aged women to a specified level. The number of reproductive-aged women living in maternity care deserts does not decrease significantly with each expanded obstetric unit. In contrast, a small number of expanded CCO services dramatically reduces the number of reproductive-aged women living further than 50 miles from CCO services.

**Figure 3.**
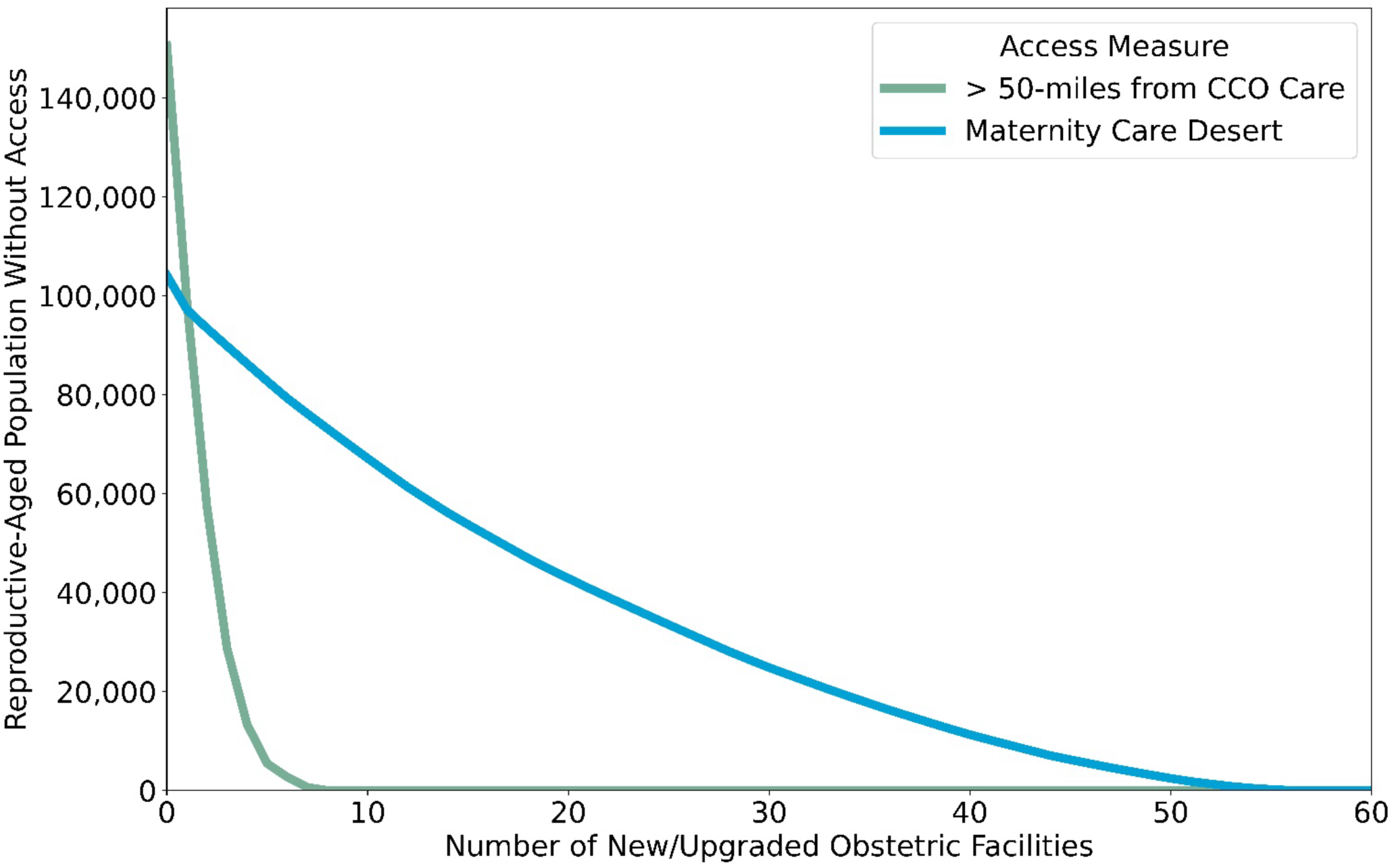
The number of obstetric care facilities needed to reduce the number of reproductive-aged (RA) women who lack access to obstetric care according to two measures of access.

## 4. Discussion

Access to care is an important dimension to consider in the context of the maternal health crisis in the U.S. Our study analyzed the implications of using existing measures of access to obstetric care as key performance indicators to evaluate and track improvements in access.

In this paper, we analyzed two current measures of obstetric access, including the popular maternity care deserts measure. Maternity care deserts are counties in which there are no obstetric providers or obstetric care facilities. This measure has been widely used in both academic literature and popular media, and it has drawn widespread attention to the lack of access to obstetric care in the U.S. Consistent with the March of Dimes report, we found that 5.5% of reproductive-aged women in Georgia live in the 56 counties designated as maternity care deserts (more than the national average, 3.5%).^15^ We found that 7.9% of reproductive-aged women live further than 50 miles from CCO services, which is less than a study using 2015 data which found that 10.2% of reproductive-aged women live further than 50 miles from CCO services.^20^ This difference may be due to a difference in distance metrics or the procedures for identifying the locations and levels of obstetric hospitals. We additionally found that 2.0% of reproductive-aged women live in regions that are both maternity care deserts and further than 50 miles from CCO services.

In our analysis, we considered the hypothetical implications of using current access measures to inform facility expansions, with the goal of evaluating these measures without concern for costs or workforce barriers. Our optimization model showed that eliminating maternity care deserts in Georgia would require at least 56 new obstetric hospitals. Doing so would increase the number of obstetric hospitals in Georgia by 67%, from 83 to 139. In contrast, ensuring all reproductive-age women in Georgia live within 50 miles of CCO services would require upgrading at least 8 existing lower-level hospitals to provide CCO services. Thus, these different measures of access imply very different strategies to expand access and very different estimates of how many obstetric facilities of different levels are needed in a geographic region.

Our findings suggest that additional tools are needed to provide estimates of how many facilities of each level of care are needed and can be sustained in a geographic region. Ideally, the number of facilities, their level of care designations, and coordination should promote optimal pregnancy outcomes. Access to obstetric care has been identified as an important opportunity to improve maternal outcomes and disparities, as rural residence has been associated with a greater probability of severe maternal morbidity and mortality,^10^ and maternity care deserts associated with higher rates of preterm birth, infant mortality, low birth weight, and maternal mortality.^16,17,37^

However, the maternity care desert measure is inherently dependent on the number and size of counties in a state and fails to account for actual distance to healthcare facilities. Counties were determined by territories and states without standardization, resulting in high variability in the number and size of counties across states.^38^ For example, Georgia has the second most counties of any state (159), only behind Texas (254), although Georgia is the 8th most populated state in the U.S. and 24th largest by area. Thus, this measure may encourage a large number of obstetric units in Georgia simply because Georgia has a large number of counties, despite the fact that 82% of reproductive-aged women who live in maternity care deserts in Georgia live within 25 miles of an obstetric hospital.

Considering these measures of access alone to inform facility expansion could lead to unintended negative consequences. We showed that it would require a 67% increase in the number of obstetric hospitals to ensure no reproductive-aged women live in maternity care deserts in Georgia. Even if the economic forces would allow for so many obstetric facilities, a maternal healthcare system with that many obstetric facilities could have unintended negative consequences due to the dilution of volume across many low-volume rural hospitals, which are known to be associated with poor pregnancy outcomes.^39–42^ Moreover, staffing this many units would likely be very expensive and challenging given that there are already obstetric workforce shortages in Georgia.^43^

While distance to CCO services could be a useful measure of access, this measure alone neither considers whether there are other nearby facilities that offer potentially sufficient lower-levels of obstetric care nor coordination between lower-level and CCO facilities. Additionally, the threshold of 50 miles to CCO services has not been validated in obstetrics,^35,36^ nor does it account for transportation factors that influence actual driving time. Thus, there are a variety of limitations in using existing measures of access alone to inform the number of facilities that are needed in a geographic region. Our findings motivate the need for nuanced access to obstetric care measures that are capable of evaluating and planning action toward the reduction of lack of access, and new approaches to estimate the optimal number of facilities of different levels of care that are necessary and sustainable within a geographic region. Future work may consider other measures of access or access expansion interventions that incorporate home visits, telemedicine, and transportation programs.

Our study is not without limitations. We use facility and population data from 2017 because the most recent publicly available data on obstetric facilities was published by the Georgia Department of Public Health in 2017. Because of the age of our data, some obstetric hospitals likely closed, opened, and merged since 2017. Specifically, the Georgia Hospital Association reports that 13 hospitals in Georgia have closed since 2013 (as of November 2022).^44^ The only obstetric hospital that closed was Wellstar Atlanta Medical Center, which closed in November 2022. This hospital was 1 mile from the Atlanta Region Perinatal Regional Center which provides CCO services. Moreover, we found that our models’ determination of maternity care deserts was consistent with the March of Dimes maternity care deserts dashboard.^33^ We expect that even with some facility closures or expansions of obstetric services at existing hospitals, our conclusion that the maternity care deserts measure is not a practical performance indicator of improvements to access to obstetric care remains. Also, we did not account for geographical barriers or traffic when calculating distance from the centroid of a census block group when computing whether the group is further than 50 miles from CCO services, and we did not account for measurement errors in the ACS. We did not account for other important barriers to access, such as transportation disadvantage and insurance coverage. We also did not account for out-of-state hospitals that offer obstetric services that could provide care to pregnant people in Georgia. Finally, our analysis only considered potential access. Future work may investigate the impact of facility expansion on realized access to care, especially considering some patients prefer to bypass local hospitals to receive care elsewhere.^45,46^

## 5. Conclusion

Our findings suggest that the current measures of obstetric access, while useful for capturing certain dimensions of the maternal healthcare system, may not be useful for estimating the optimal number, designations, and coordination of obstetric care within a geographic region. Specifically, while maternity care deserts are associated with increased rates of maternal mortality,^16^ this measure is not a practical performance indicator of improvements to access to obstetric care. Thus, there is a need for tools that can track improvements and inform the appropriate number of obstetric care facilities that are needed in a geographic region to improve access to high-quality, risk-appropriate care, and ultimately improve obstetric outcomes. In addition, future work may examine how to optimally balance the cost and outcomes of expanding care, considering the trade-offs between increased access and loss of quality due to dilution and staffing issues, and incorporating alternate access expansion strategies such as home visits, telemedicine, and transportation programs.

## Declarations

### Ethics approval and consent to participate

The Georgia Institute of Technology Internal Review Board deemed this study qualified for a waiver of consent.

### Consent for publication

Not applicable.

### Availability of data and materials

Georgia hospital data is available in public records from Georgia’s Department of Public Health from 2017.^27^ All Georgia block group population counts data are publicly available from the U.S. Census Bureau. Link: https://data.census.gov/all?q=acs&g=040XX00US13$1500000&y=2017. Georgia block group population centroids are publicly available from the U.S. Census Bureau. Link: https://www.census.gov/geographies/reference-files/time-series/geo/centers-population.2010.html#list-tab-ZWAU50627XERV1TT2V. March of Dimes maternity care desert data is available in the March of Dimes report^15^ and the March of Dimes maternity care deserts dashboard. Link: https://www2.deloitte.com/us/en/pages/life-sciences-and-health-care/articles/march-of-dimes-maternity-care-deserts-dashboard.html.

### Authors’ contributions

MEM collected the data, used programming to conduct the analysis, and prepared all figures and tables. LNS and MEM developed the methodology and wrote the manuscript text. SMR contributed domain knowledge and reviewed and edited the manuscript.

## Data Availability

Code used in the the present study are available upon reasonable request to the authors. Georgia hospital data is available in public records from Georgia’s Department of Public Health from 2017. (See reference 27) All Georgia block group population counts data are publicly available from the U.S. Census Bureau. Link: https://data.census.gov/all?q=acs&g=040XX00US13$1500000&y=2017. Georgia block group population centroids are publicly available from the U.S. Census Bureau. Link: https://www.census.gov/geographies/reference-files/time-series/geo/centers-population.2010.html#list-tab-ZWAU50627XERV1TT2V. March of Dimes maternity care desert data is available in the March of Dimes report (see reference 15) and the March of Dimes maternity care deserts dashboard. Link: https://www2.deloitte.com/us/en/pages/life-sciences-and-health-care/articles/march-of-dimes-maternity-care-deserts-dashboard.html.

## Acknowledgements

The authors would like to thank Hengyi Hu and Abel Sapirstein for their assistance with the data collection and Dr. Debra Kane for her thoughtful comments throughout the analysis.

## Appendix: Mathematical optimization models

In this appendix, we present our mathematical optimization models used to compare facility expansion policies. This decision-analytic approach provides a way to optimally allocate resources (e.g., facility expansions) across a system in a way that considers constraints (e.g., no more than 4 facilities can be expanded) and decision makers’ objectives (e.g., minimize the number of reproductive-aged (RA) women living in deserts).

### Minimize the number of reproductive-aged women who live in maternity care deserts

We consider a set of counties *C* = {1,2, …, *C*} and a set of census block groups ℬ = {1,2, …, *B*}. The population of reproductive-aged women in census block group *b* is denoted by *p_b_*. We use an indicator β*_bc_* which takes on a value of 1 if census block group *b* is within a county *c.* We use an indicator *q_c_* which takes on a value of 1 if county *c* has no obstetric providers practicing within the county. We use an indicator 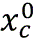 which takes on a value of 1 if county *c* has no hospital or birth center providing obstetric care within the county.

For a county to be considered a maternity care desert, it must have no obstetric providers practicing and no hospital or birth center providing obstetric care within the county.

We consider whether a county is a maternity care desert or not using an indicator *d_c_*, which takes of a value of 1 if a county *c* is a maternity care desert and 0 otherwise. If a county is a maternity care desert, then all block groups within the county are also maternity care deserts. We indicate if a block group is a maternity care desert using an indicator *d_b_*,which takes on a value of 1 if census block group *b* is a desert and 0 otherwise. Thus, the total number of reproductive-aged women who currently live in maternity care deserts is given by: 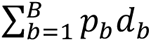.

To consider the impact of policy interventions, we consider the possibility that facilities can be expanded to provide maternity care. We designate *decision variables* 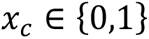 such that 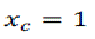 indicates county *c* has a hospital or birth center providing maternity care after infrastructure is expanded. We consider a *constraint* that we can expand at most *F* facilities to provide maternity care: 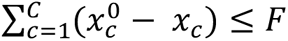. We also consider a set of *constraints* that each county is considered a desert if it has no obstetric providers and no hospital or birth centers within the county: 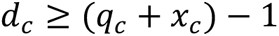. Further, we add another set of *constraints* that each block group is considered a desert only if it is within a county that is a desert: 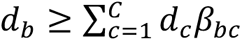. Finally, we add a set of *constraints* 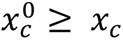 which enforce that no county can be downgraded such that they no longer have a hospital or birth center providing obstetric services.

Our optimization model will select the values of the decision variables that satisfy the constraints in order to minimize our objective function. To minimize the total number of reproductive-aged women in maternity care deserts, we will minimize the following objective function:

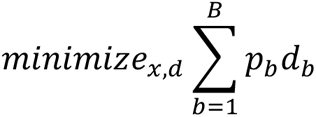

Thus, our final optimization model is:

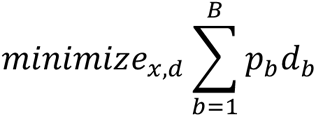

Subject to:

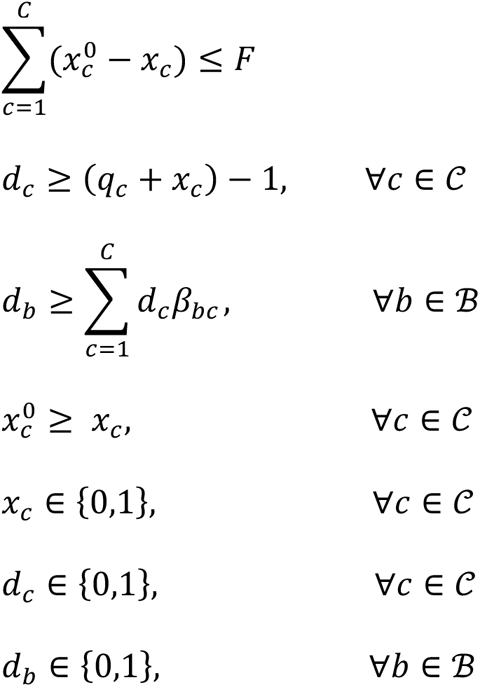

We can solve this problem quickly by ranking the counties in terms of *p*_*c*_ and selecting the *F* largest values. We solve this problem for a range of *F* values (*F* = 1,2,3,…) to evaluate how the number of reproductive-aged women living in maternity care deserts changes.

### Minimize the number of women of who live more than 50 miles from CCO services

We consider a set of census block groups ℬ = {1,2, …, *B* }, and the population of reproductive-aged women in census block group *b* is denoted by *p_b_*. We also consider a set of obstetric facilities ℋ = {1,2, …, *H*}. We use an indicator 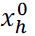 which takes on a value of 1 if facility *h* provides CCO services. We use an indicator α*_bh_* which takes on a value of 1 if census block group *b* is within 50 miles of facility *h*.

We consider whether a census block group is further than 50 miles from CCO services using an indicator *d_b_*, which takes of a value of 1 if a census block group *b* is further than 50 miles from CCO services and 0 otherwise. Thus, the total number of women who live further than 50 miles from CCO services is given by: 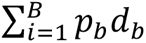.

To consider the impact of policy interventions, we allow lower-level facilities that do not provide CCO services to be upgraded to provide CCO services. We consider *decision variables x_h_* ∈ {0,1} such that *x_h_* = 1 indicates that facility *h* provides CCO services after upgrades. We consider a *constraint* that we can upgrade at most *F* facilities to provide critical care obstetric services: 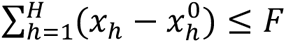. Further, we add a set of constraints 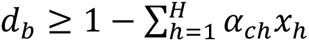 which enforce that a census block group lacks access if it is further than 50 miles from its nearest obstetric facility offering CCO services. Finally, we add a set of constraints 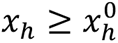 which enforce that no facilities can be downgraded such that they no longer provide CCO services.

Our optimization model will select the values of the decision variables that satisfy the constraints in order to minimize our objective function. To minimize the total number of reproductive-aged women who live further than 50 miles to CCO services, we will minimize the following objective function:

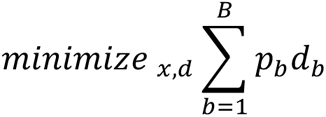

Thus, our final optimization model is:

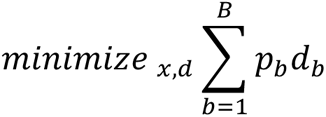

Subject to:

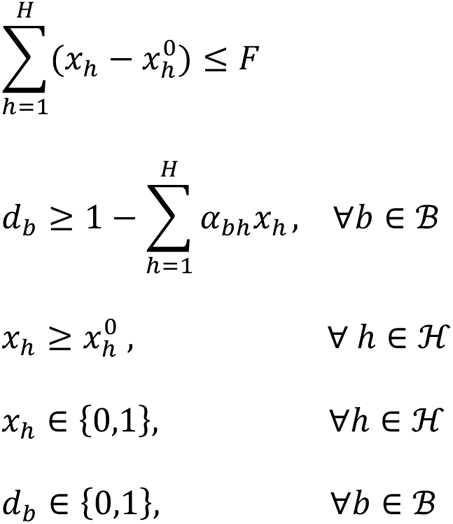

We solve this problem for a range of *F* values.

